# Delta variant and mRNA Covid-19 vaccines effectiveness: higher odds of vaccine infection breakthroughs

**DOI:** 10.1101/2021.08.14.21262020

**Authors:** Irina Kislaya, Eduardo Freire Rodrigues, Vítor Borges, João Paulo Gomes, Carlos Sousa, José Pedro Almeida, André Peralta-Santos, Baltazar Nunes, on behalf of PT-Covid-19 Group

## Abstract

**Background:** The SARS-CoV-2 Delta variant (B.1.617.2), initially identified in India, has become predominant in several countries, including Portugal. Few studies have compared the effectiveness of mRNA vaccines against Delta versus Alpha variant of concern (VOC) and estimated variant-specific viral loads in vaccine infection breakthroughs cases. In the context of Delta dominance, this information is critical to inform decision-makers regarding the planning of restrictions and vaccination roll-out.

**Methods:** We developed a case-case study to compare mRNA vaccines’ effectiveness against Delta (B.1.617.2) versus Alpha (B.1.1.7) variants. We used RT-PCR positive cases notified to the National Surveillance System between 17th of May and 4th of July 2021 (week 20 to 26) and information about demographics and vaccination status through the electronic vaccination register. Whole-genome sequencing (WGS) or spike (S) gene target failure (SGTF) data were used to classify SARS-CoV-2 variants. The odds of vaccinated individuals to become infected (odds of vaccine infection breakthrough) in Delta cases compared to Alpha SARS-CoV-2 cases was estimated by conditional logistic regression adjusted for age group, sex, and matched by the week of diagnosis. As a surrogate of viral load, mean RT-PCR Ct values were stratified and compared between vaccine status and VOC.

**Results:** Of the 2 097 SARS-CoV-2 RT-PCR positive cases included in the analysis, 966 (46.1%) were classified with WGS and 1131 (53.9%) with SGTF. Individuals infected with the Delta variant were more frequently vaccinated 162 (12%) than individuals infected with the Alpha variant 38 (5%). We report a statistically significant higher odds of vaccine infection breakthrough for partial (OR=1.70; CI95% 1.18 to 2.47) and complete vaccination (OR=1.96; CI95% 1.22 to 3.14) in the Delta cases when compared to the Alpha cases, suggesting lower mRNA vaccine effectiveness against Delta cases. On our secondary analysis, we observed lower mean Ct values for the Delta VOC cases versus Alpha, regardless the vaccination status. Additionally, the Delta variant cases revealed a Ct-value mean increase of 2.24 (CI95% 0.85 to 3.64) between unvaccinated and fully vaccinated breakthrough cases contrasting with 4.49 (CI95% 2.07 to 6.91) in the Alpha VOC, suggesting a lower impact of vaccine on viral load of Delta cases.

**Conclusions:** We found significantly higher odds of vaccine infection breakthrough in Delta cases when compared to Alpha cases, suggesting lower effectiveness of the mRNA vaccines in preventing infection with the Delta variant. Additionally, the vaccine breakthrough cases are estimated to be of higher mean Ct values, suggesting higher infectiousness with the Delta variant infection. These findings can help decision-makers weigh on the application or lifting of control measures and adjusting vaccine roll-out depending on the predominance of the Delta variant and the coverage of partial and complete mRNA vaccination.

## Background

The SARS-CoV-2 B.1.617.2 lineage, also known as Delta Variant of Concern (VOC), first sequenced in India in December 2020, was identified in Portugal in late April and quickly became dominant, reaching 90% of all sequenced cases in late June 2021 (week 26), just two months after firstly identified [1]. Available evidence suggests that this VOC is associated with higher transmissibility, risk of hospitalization and reduced antibody neutralization compared to other VOCs [2,3].

Vaccination is the primary pharmacological measure to control the transmission of SARS-CoV-2 and mitigate its impact on hospitalizations and mortality. In Portugal, vaccination was initiated in late December 2020 for those at higher risk of severe disease or exposure and has since February 2021 been rolled out by descendent age criteria. By week 26 (28^th^ June to 4^th^ July), 36% of the population was fully vaccinated, and 56% had started, or completed vaccination (Figure S1), the majority (75%) with mRNA vaccines (BNT162b2 (*Comirnaty*) or mRNA-1273 SARS-CoV-2 (*Covid-19 Vaccine Moderna*)) administrated with a 28-day dose interval [4].

Early reports of vaccine effectiveness indicate a high protection of mRNA vaccines against infection and disease [5,6] and a reduced viral load in the vaccinated cases [7,8]. However, reports of vaccine effectiveness against Delta have shown a decreased protection of the vaccines compared to the Alpha variant [2,9]. Validating this potential reduction of the vaccine effectiveness against the Delta VOC is critical to inform further public health measures, particularly as it becomes globally dominant. This study aimed to provide a measure of compared effectiveness of mRNA vaccines (BNT162b2 (*Comirnaty*) and mRNA-1273 SARS-CoV-2 (*Covid-19 Vaccine Moderna*)) against B.1.617.2 (Delta) versus B.1.1.7 (Alpha) VOC, using a case-case study design. As a secondary objective the RT-PCR cycle threshold values (Ct values) between vaccine status for Alpha and Delta variants were compared as an indirect measure of viral load and thus transmissibility of the vaccine breakthrough cases for both variants.

## Methods

### Study design

We developed an observational case-case study [10] comparing odds of vaccination between reverse transcription polymerase chain reaction (RT-PCR) positive cases (symptomatic or asymptomatic) classified as Delta versus Alpha VOC. The study period was defined between the 17th of May and the 4th of July 2021 (respectively, week 20 and week 26, inclusive), to cover the period of VOC replacement in Portugal, from the Alpha (84.8%, week 19) to Delta dominance (96.1%, week 27)[1]. Our analysis included individuals with data on whole-genome sequencing (WGS) or spike (S) gene target failure (SGTF), aged 40 or more years old, eligible for vaccination in the study period. Individuals with missing data on national health registry number, age, sex, or diagnosis date, and those vaccinated with Ad26.COV2-S (*Covid-19 Vaccine Janssen*) or ChAdOx1 nCoV-19 (*Vaxzevria*) vaccines were excluded from the study.

In order to indirectly infer the level of infectiousness of cases according to vaccination status and VOC type, a secondary analysis was performed by comparing the paired means of RT-PCR Ct values for nucleocapsid (N) and open reading frame 1ab (ORF1ab) genes. Lower Ct values reflect a reduced number of PCR cycles required for amplification of SARS-CoV-2 RNA and, therefore, a higher number of virus copies within the sample. As such, studies have used Ct values to estimate viral load or viral shedding [8,11].

### Data Sources

#### SARS-CoV-2 Cases

RT-PCR testing for SARS-CoV-2 in Portugal is done by hospitals as well as by public and private laboratories, and is available free of charge to anyone with symptoms consistent with Covid-19[12]. Laboratory confirmed cases are notified to the mandatory National Epidemiological Surveillance Information System (SINAVE). For this study, each notifying laboratory selected a random sample of PCR-positive nasopharyngeal samples collected between the 17^th^ of May and the 4^th^ of July 2021 (respectively, week 20 and week 26) to be sent to the National SARS-CoV-2 Genomic Surveillance Network [1,13] and, thus, to be included in the study. We also included samples from a private molecular biology laboratory (UNILABS) with nationwide coverage that routinely performs analysis on SGTF. Ct values for RT-PCR were collected as semi-quantitative measure to categorize viral load [11]. Duplicated records were removed based on national health register numbers, maintaining only the first collected sample.

### Variant classification

SARS-CoV-2 variants were classified by viral whole-genome sequencing (WGS) or inferred by spike (S) gene target failure (SGTF) data. For non-sequenced samples, S-positive specimens (with no amplification of S gene) were considered Delta, while SGTF were considered Alpha, using the TaqPath™ Covid 19 CE IVD RT-PCR Kit (Thermo Scientific™) assay that targets three gene: S, N, and ORF1ab, performed according to the manufacturer’s specifications as described elsewhere [13].

### Vaccination status, demographics, and data linkage

Covid-19 vaccination status was obtained through the electronic national vaccination register (VACINAS). Vaccination exposure was classified as: (i) no register of vaccine administration prior to diagnosis (unvaccinated); (ii) SARS-CoV-2 infection diagnosis less than 14 days after first dose mRNA vaccine uptake (1 dose | less 14 days); (iii) SARS-CoV-2 infection diagnosis 14 or more days after first dose uptake or less than 14 days after second dose uptake (1 dose | more 14 days or 2 doses | less 14 days; partial vaccination); and (iv) 14 or more days following second dose mRNA vaccine uptake (2 doses | more 14; complete vaccination). Information about age, sex and date at diagnosis was routinely collected by the SINAVE. Data linkage between all data sources was conducted using the national health register number.

### Statistical Analysis

Characteristics of Delta and Alpha SARS-CoV-2 cases were compared using the chi-square test. Delta cases were considered as cases of interest, while Alpha cases were considered as the reference group. Conditional logistic regression matched by the week of diagnosis and adjusted for age group and sex was used to estimate confounder-adjusted odds of having been infected by SARS-CoV-2 and vaccinated in Delta cases compared to Alpha SARS-CoV-2 cases. The abovementioned covariates might be associated with the probability of having been vaccinated and being exposed to the virus and type of variant.

In our analysis, an odds ratio (OR) equal to one (OR=1) indicates no difference in odds of having been infected by SARS-CoV-2 and vaccinated and, thus, a proxy of no difference between mRNA vaccine effectiveness against Delta versus Alpha variant. An odds ratio above one (OR>1) indicates a higher odds of having been infected by SARS-CoV-2 and vaccinated, as such, lower vaccine effectiveness against Delta in comparison with Alpha, while an odds-ratio below one (OR<1) indicates a lower odds of having been infected by SARS-CoV-2 and vaccinated among Delta cases and respectively a higher vaccine effectiveness against Delta in comparison to Alpha VOC (more information in the supplementary material).

Mean and standard deviation (SD) Ct values for Alpha and Delta cases were stratified according to vaccination status. Differences between mean Ct values by vaccination status and VOC were evaluated by fitting a linear multiple regression model with Ct values as outcome adjusting for sex, age group and week of case diagnosis. An interaction term between vaccination status and VOC type was included in the regression model to measure if vaccination status effect on Ct values is differential between Delta and Alpha VOC.

### Sensitivity analysis

To assess the change of the sampling strategy for WGS from a monthly to weekly basis, which occurred on week 21, we restricted our analysis to weeks 22-26. Additionally, to assess the bias of misclassification error associated with the SGTF method — particularly in the early weeks of the study period where the overall prevalence of the Delta variant was lower, and possibly SGTF sensitivity was also lower — we analyzed samples identified exclusively through WGS during weeks 22-26. Finally, to address if having been infected and vaccinated was associated with lower infectiousness in any of the studied VOCs, we restricted analysis to the samples for Ct values below 25 (Ct<25)[8].

## Results

### Main analysis

Overall, 2 097 SARS-CoV-2 PCR positive cases were included in the analysis; of those, 966 (46.1%) were variant classified with WGS and 1131 (53.9%) with SGTF. During the study period, 94.7% (827/873) of the S-positive sequenced cases were confirmed as Delta, and 96.9% (372/384) of SGTF samples were classified as Alpha through WGS, thus indicating that the SGTF-derived VOC classification was robust.

Among Delta cases, we observed a higher proportion of individuals older than 70 years (p<0.001) (Table 1), and a higher proportion of vaccinated individuals (p<0.001) when compared with the Alpha cases.

**Table 1.** Characteristics of the study sample by variant of concern

|  | **Delta (B.1.617.2)** | **Alpha (B.1.1.7)** | **Total** |
| --- | --- | --- | --- |
|  | n(%) | n(%) | n(%) |
| Overall | 1366 | 731 | 2097 |
| **Week of diagnosis (2021)** |  |  |  |
| 20 | 53 (4) | 137 (19) | 190 (9) |
| 21 | 64 (5) | 154 (21) | 218 (10) |
| 22 | 112 (8) | 171 (23) | 283 (13) |
| 23 | 192 (14) | 114 (16) | 306 (15) |
| 24 | 249 (18) | 73 (10) | 322 (15) |
| 25 | 350 (26) | 38 (5) | 388 (19) |
| 26 | 346 (25) | 44 (6) | 390 (19) |
| **Age group** |  |  |  |
| 40-49 | 760 (56) | 378 (52) | 1138 (54) |
| 50-69 | 474 (35) | 313 (43) | 787 (38) |
| 70+ | 132 (10) | 40 (5) | 172 (8) |
| **Sex** |  |  |  |
| Female | 707 (52) | 402 (55) | 1109 (53) |
| Male | 659 (48) | 329 (45) | 988 (47) |

**Table 2.**
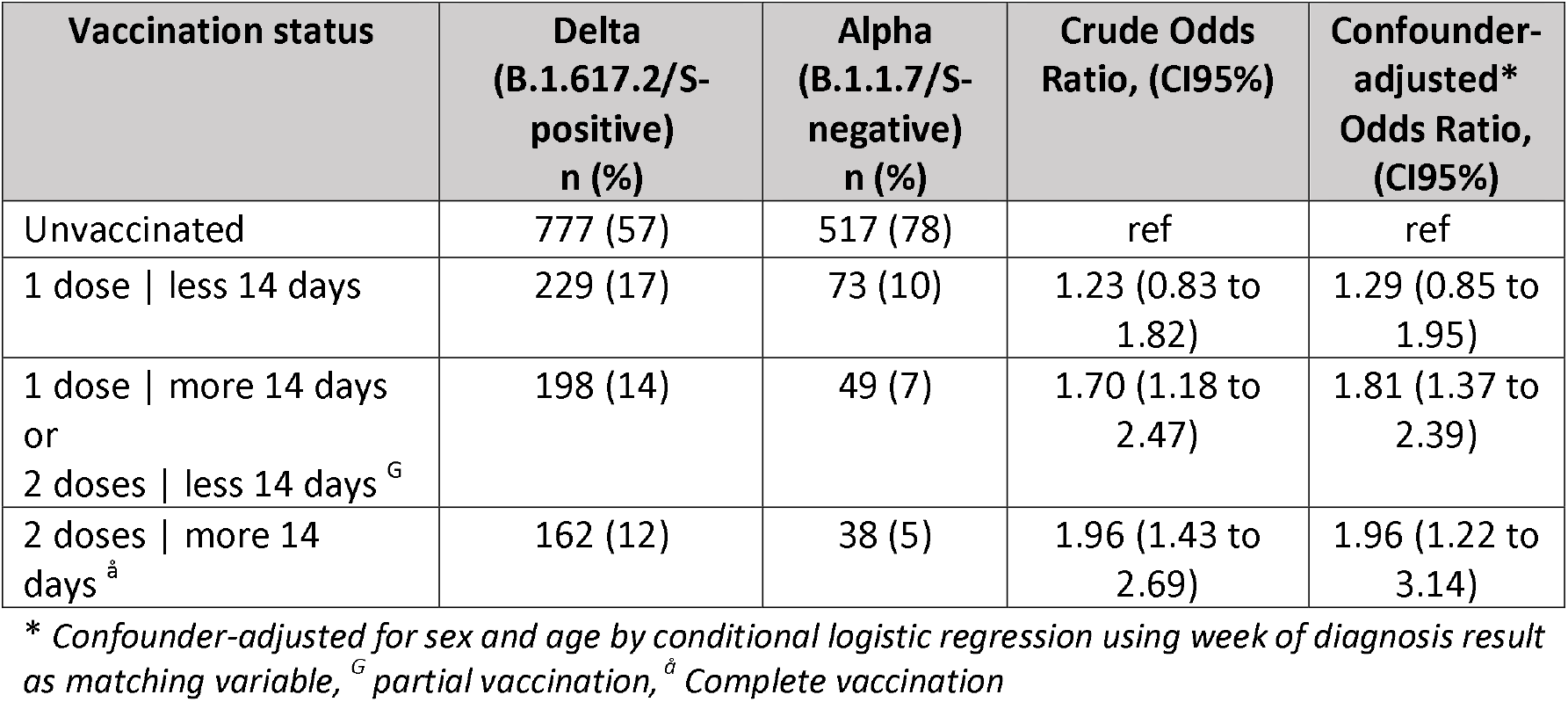
Crude and adjusted odds ratio of being infected and vaccinated (odds of vaccine infection breakthrough) in Delta cases compared to Alpha SARS-CoV-2 cases

| **Vaccination status** | **Delta**<br>**(B.1.617.2/S-**<br>**positive)**<br>**n (%)** | **Alpha**<br>**(B.1.1.7/S-**<br>**negative)**<br>**n (%)** | **Crude Odds**<br>**Ratio, (CI95%)** | **Confounder-**<br>**adjusted\***<br>**Odds Ratio,**<br>**(CI95%)** |
| --- | --- | --- | --- | --- |
| Unvaccinated | 777 (57) | 517 (78) | ref | ref |
| 1 dose \| less 14 days | 229 (17) | 73 (10) | 1.23 (0.83 to 1.82) | 1.29 (0.85 to 1.95) |
| 1 dose \| more 14 days<br>or<br>2 doses \| less 14 days <sup>G</sup> | 198 (14) | 49 (7) | 1.70 (1.18 to 2.47) | 1.81 (1.37 to 2.39) |
| 2 doses \| more 14 days <sup>A</sup> | 162 (12) | 38 (5) | 1.96 (1.43 to 2.69) | 1.96 (1.22 to 3.14) |

\* Confounder-adjusted for sex and age by conditional logistic regression using week of diagnosis result as matching variable, <sup>G</sup> partial vaccination, <sup>A</sup> Complete vaccination

We report a statistically significant higher odds of being partially vaccinated (OR=1.70; CI95% 1.18 to 2.47) and completely vaccinated (OR=1.96; CI95% 1.22 to 3.14) in the Delta cases when compared to the Alpha cases, suggesting lower mRNA vaccine effectiveness for the Delta variant.

After adjustment for age group and sex, no changes in point estimate were observed for complete vaccination scheme (OR=1.96; CI95% 1.43 to 2.69), while for partial vaccination (14 or more days after first dose uptake), the OR point estimate presented a small increase (OR=1.81; CI95% 1.37 to 2.39).

### Secondary analysis

Comparing cases by vaccination status, we observed higher mean Ct values among those with complete vaccination scheme relative to unvaccinated for both VOC, 17.7 vs. 16.6 for Delta and 21.8 vs. 18.4 for Alpha, respectively (Table 3, Figure 1), suggestive of lower viral loads in vaccinated compared to unvaccinated cases. While the Alpha variant cases had a Ct values mean difference (MD) of 4.49 (CI95% 2.07 to 6.91) after complete vaccination, representing an increase of Ct values (lower infectiousness), the Delta variant breakthrough cases presented only about half of that increase, with a Ct value statistically significant mean difference point estimate of 2.24 (95% CI: 0.85 to 3.64) between unvaccinated and fully vaccinated breakthrough cases.

**Table 3.**
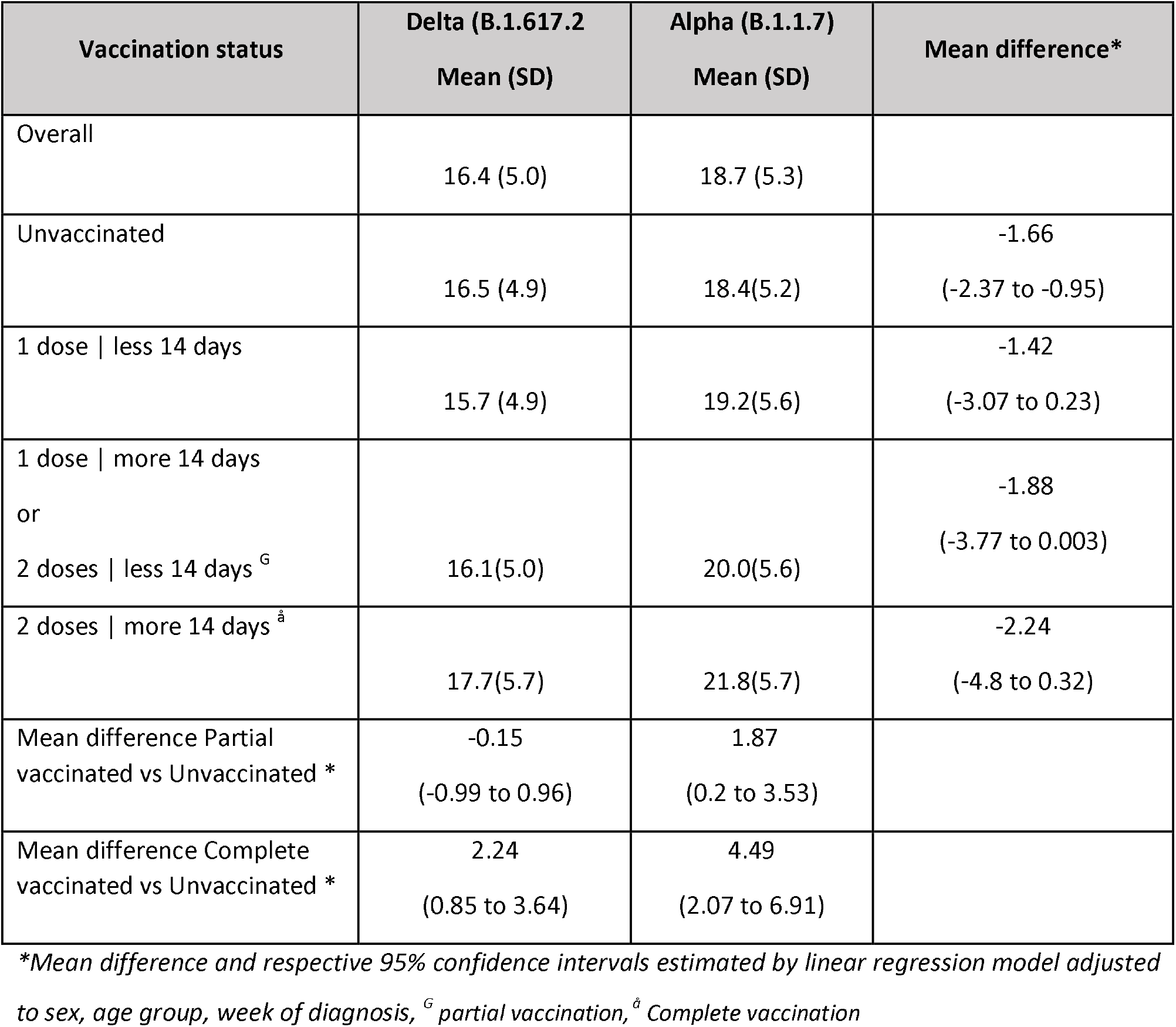
Mean and standard deviation Ct values (mean N and ORF gene) stratified by vaccination status and Delta or Alpha variant

**Table 4.** Sensitivity analysis between week 22 and 26, n=1689, adjusted odds ratio of being infected and vaccinated (odds of vaccine infection breakthrough) in Delta cases compared to Alpha SARS-CoV-2 cases

| Vaccination status | Delta (B.1.617.2)<br>n (%) | Alpha (B.1.1.7)<br>n (%) | Crude Odds Ratio<br>(CI95%) | Confounder-adjusted* Odds<br>Ratio (CI95%) |
| --- | --- | --- | --- | --- |
| Restricted to<br>weeks 22-26, n=1689 |  |  |  |  |
| Unvaccinated | 682 (55) | 328 (75) | ref | ref |
| 1 dose \| less 14 days | 224 (18) | 54 (12) | 1.34<br>(0.94 to 1.91) | 1.38<br>(0.96 to 1.98) |
| 1 dose \| more 14 days<br>or 2 doses \| less 14 days | 190 (15) | 32 (7) | 1.87<br>(1.22 to 2.86) | 1.91<br>(1.22 to 3.00) |
| 2 doses \| more 14 days<sup>â</sup> | 153 (12) | 26 (6) | 1.99<br>(1.25 to 3.16) | 1.78<br>(1.01 to 3.13) |
| Restricted to WGS<br>classified VOC cases,<br>n=931 |  |  |  |  |
| Unvaccinated | 406 (59) | 189 (77) | ref | ref |
| 1 dose \| less 14 days | 104 (15) | 29 (12) | 1.21<br>(0.75 to 1.95) | 1.20<br>(0.73 to 1.96) |
| 1 dose \| more 14 days<br>or 2 doses \| less 14 days | 84 (12) | 11 (4) | 2.63<br>(1.34 to 5.20) | 2.50<br>(1.23 to 5.08) |
| 2 doses \| more 14 days<sup>â</sup> | 90 (13) | 18 (7) | 1.91<br>(1.09 to 3.34) | 1.48<br>(0.75 to 2.93) |
| Restricted to cases<br>with Ct value < 25,<br>n=1363 |  |  |  |  |
| Unvaccinated | 492 (57) | 412 (82) | ref | ref |
| 1 dose \| less 14 days | 161 (19) | 48 (10) | 1.20<br>(0.79 to 1.81) | 1.30<br>(0.85 to 1.98) |
| 1 dose \| more 14 days<br>or 2 doses \| less 14 days | 142 (17) | 30 (6) | 2.14<br>(1.32 to 3.46) | 2.47<br>(1.48 to 4.12) |
| 2 doses \| more 14 days<sup>â</sup> | 64 (7) | 14 (3) | 1.92<br>(0.97 to 3.82) | 2.42<br>(1.06 to 5.51) |

\* Confounder-adjusted for sex and age by conditional logistic regression using week of diagnosis result as matching variable, <sup>g</sup> partial vaccination, <sup>â</sup> Complete vaccination

**Figure 1 –.**
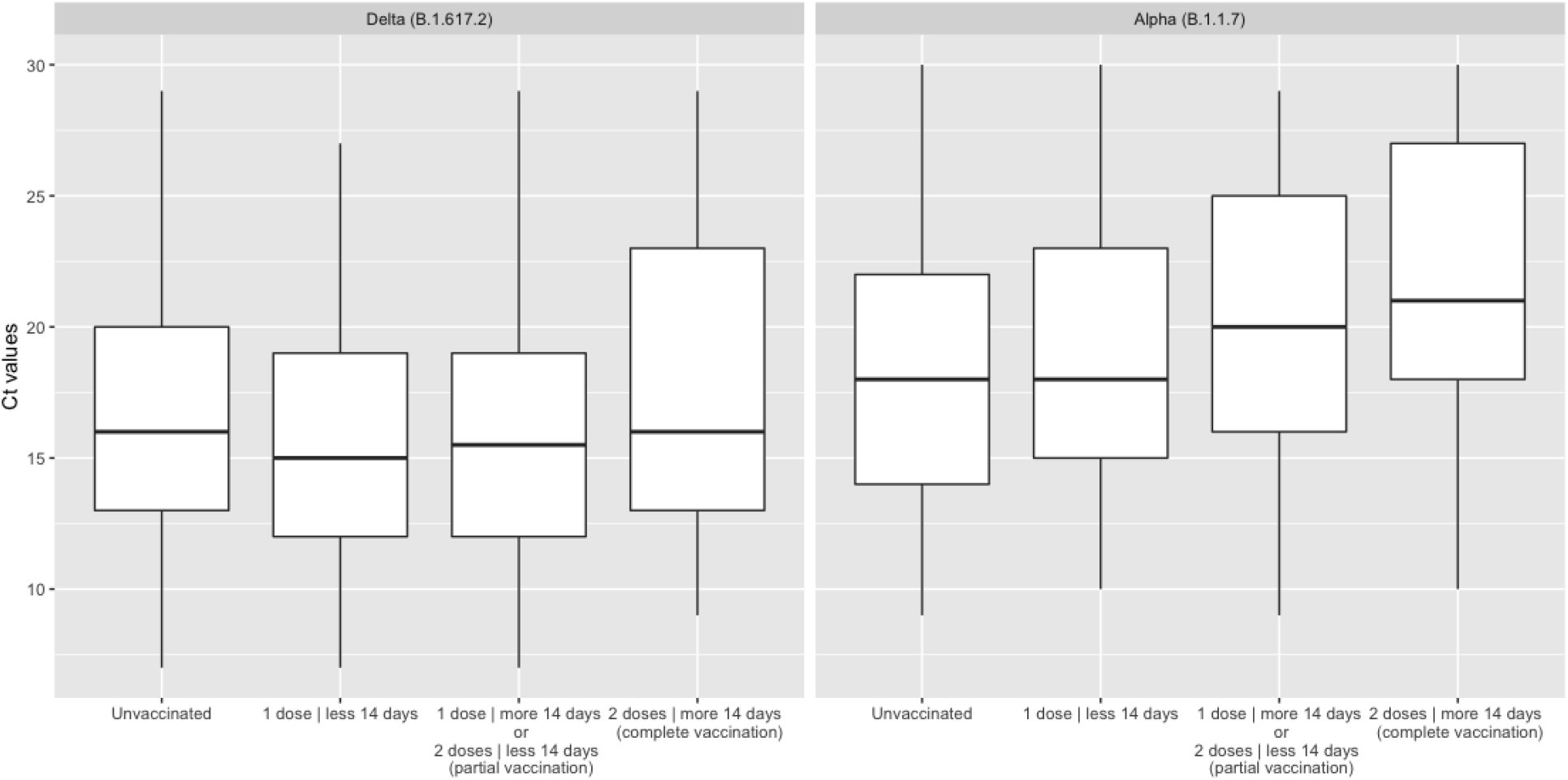
Ct value paired means distribution (Ct N and Ct ORF1ab) by variant and vaccination status

For partial vaccination, statistically significant differences in mean Ct values were observed for Alpha (MD=1.87; CI95%0.2 to 3.53), but not for Delta cases (MD=-0.15; CI95% -0.99 to 0.96), suggesting similar viral load between unvaccinated and partial vaccinated Delta cases.

In secondary analysis by VOC regardless the vaccination status, lower mean Ct values were observed for the Delta VOC cases versus Alpha (16.4 vs.18.7). The confounder-adjusted paired-mean difference in Ct values between two VOCs was also statistically significant for unvaccinated (MD= -1.66; CI95% -2.37 to - 0.95), while for partial (MD=–1.88; CI95% -3.77 to 0.003) and complete vaccination (MD= -2.24; CI95% - 4.8 to 0.32) we observed mean differences between two VOCs without statistical significance.

### Sensitivity analysis

In a sensitivity analysis, restricting to weeks 22 to 26, confounder-adjusted OR estimates for having been infected and complete vaccinated (OR=1.78; CI95% 1.01 to 3.13) and partial vaccination (OR=1.91; CI95% 1.22 to 3.00) remained similar and statistically significant. Restricting the analysis to the cases identified through WGS, we observed a drop in the adjusted OR point estimate of complete vaccination (OR=1.48; CI95% 0.75 to 2.93) with a loss of statistical significance, but for partial vaccination, estimates yielded increased significant results (OR= 2.5; CI95% 1.23 to 5.08). Restricting to cases with Ct value < 25 (n=1363), an increase of the OR of infection and vaccination against Delta versus Alpha variants was observed both for partial (OR=2.47; CI95% 1.48 to 4.12) and complete vaccination (OR= 2.42; CI95% 1.06 to 5.51).

## Discussion and Conclusions

We observed statistically significant higher odds of having been infected and vaccinated (vaccine infection breakthrough) among Delta compared to Alpha cases, suggesting a lower mRNA vaccines effectiveness for SARS-CoV-2 infection with the Delta VOC. The findings were consistent for both complete and partial vaccination. Delta breakthrough cases have a higher viral load (lower Ct values) compared Alpha breakthrough cases.

Our results are robust to variations in the sampling strategy, including changing the weeks of diagnosis included, and the potential misclassification of using SGTF to identify Delta Variant. Additionally, considering only cases with a high potential infectiousness (Ct value < 25), we observed a noteworthy higher reduction of mRNA vaccine effectiveness against Delta versus Alpha variant.

The estimates regarding the odds ratio for complete vaccination (vaccine infection breakthrough), OR=1.96, are in line with findings of recent test-negative design (TND) studies on vaccine effectiveness from Scotland and England [2,9] based only on SGTF or mixed SGTF/WGS methodology for variants identification that reported 5.9 to 13.0 percentage points reduction of BNT162b2 vaccine effectiveness against Delta VOC compared to Alpha VOC for complete vaccination. Although the authors did not formally compare the vaccine effectiveness estimates against Alpha and Delta variants, based on non-overlapping vaccine effectiveness confidence intervals limits, it can be concluded that differences were statistically significant equivalent to case-case OR estimates ranging between 1.9 and 2.6.

For partial vaccination, our results, both based on identification through WGS (OR=2.5) or SGTF/WGS (OR=1.8), indicated statistically significant lower mRNA vaccine effectiveness against Delta VOC, supporting the need to promptly complete vaccination schemes to account for variant swiftly reduced effectiveness. Comparison to other studies should be made with prudence due to different operational definitions of exposure to the partial vaccination (Portugal: fourteen days after first dose uptake; England: twenty-one days after first dose uptake; Scotland: twenty-eight days after first dose uptake).

The first study published on the vaccine effectiveness against the Delta variant was conducted in England [9] and estimated differences in vaccine effectiveness against symptomatic infection between Delta and Alpha cases higher in magnitude (OR=2.42; CI95% 1.47 to 3.98) for complete vaccination and no differences for partial vaccination (OR=1.17; CI95% 0.88 to 1.54)[9]. Several factors may explain these differences between the abovementioned study and our work: (1) the target population is different (England 16+ vs. Portugal 40+), (2) there are also differences in the variants circulation during the study period, (3) differences in vaccination uptake, with England presenting a higher proportion of individuals exposed to two doses, (4) sample selection methodology, (5) differences in exposure definitions and time of exposure to the first dose (interval between doses recommended in England is different than in PT).

Case-case design has proven to be helpful to compare vaccine effectiveness for Covid-19 VOC due to its quick implementation and valuable insights, particularly in the context of frequent and swift VOC emergence. This study has, however, limitations. We report odds of vaccine breakthrough between Delta and Alpha VOC, which can be interpreted as a measure of the relative vaccine effectiveness. Although a case-case study design does not provide a direct measure of effectiveness against a specific VOC, it may provide substantial evidence for further public health measures to control the transmission of SARS-CoV-2 in the context of the global lifting of restrictions.

Our results include infected individuals (both asymptomatic and symptomatic) and thus cannot estimate effectiveness against symptomatic disease. However, by restricting our main analysis to samples with higher viral loads — selected based on low Ct values (Ct<25) — an even higher OR of vaccination between Delta and Alpha variant was obtained which supports the relevance of the Delta relative vaccine effectiveness reduction effect in the transmission of SARS-CoV-2, both for partial (OR=2.47; CI95%: 1.48 to 4.12) and complete vaccination (OR= 2.42; CI95% 1.06 to 5.51).

On our secondary analysis, we observed lower Ct values — indicative of higher viral loads — among Delta cases compared to Alpha after both complete (MD=-2.24; CI95% -4.8 to 0.32) and partial vaccination (MD=–1.88; CI95% -3.77 to 0.003). Furthermore, while complete vaccination increases Ct values (thus reducing estimated viral loads) on Alpha cases by 4.49 (CI95% 2.07 to 6.91), with the Delta variant case complete vaccination had a much discrete increase of Ct values 2.24 (CI95% 0.85 to 3.64), only half of the difference observed for Alpha cases. These findings are consistent with vaccine infection breakthrough cases with Delta variant having higher infectiousness than cases with Alpha. Our findings were similar to results of the studies performed in Israel, when Alpha variant was the dominant, that also found an increase of N gene Ct values of 5.09 (CI95% 2.8 to 7.4)[8] between complete vaccinated and unvaccinated cases and of 1.51 for partially vaccinated with BNT162b2[7] which is consistent with our results for the Alpha variant.

Results must always be interpreted in context, as they do not provide evidence to question the benefits of the mRNA vaccines to individual health, such as reducing symptoms, disease severity, or the impact on health services capacity.

The selection of positive cases for WGS and SGTF identification was independent from the VOC type and from vaccination status, so the probability of differential selection bias is low. Overall, 58 458 cases were identified in Portugal during the study period, 22 784 of those were older than 40 years and their age distribution was different from study sample (Figure S4) with a discrete skewness towards younger ages in the study sample. This may be explained by the fact that our sample was collected mainly through ambulatory laboratories while older (80+) individuals are expected to be more frequently diagnosed by hospital laboratories. This result could bias our estimates if the reduction of vaccine effectiveness between Delta and Alpha variant is age dependent. By example, if the vaccine effectiveness reduction is higher among older individuals our results could be underestimated.

Moreover, Delta cases had a higher proportion of older participants, and older participants were the first to be vaccinated in Portugal [4] and, thus, have a longer exposure time since 2^nd^ dose. With time, there can be a waning of the vaccine effect; this may have also contributed to the observed differences for a complete vaccination scheme.

Although we have observed a high positive predictive value (94.7%) of non-SGTF data to identify Delta cases within the study sample, some misclassification bias may lead to underestimating the reported effect. SGTF methodology had previously shown good classification accuracy to identify B.1.1.7 in Portugal [13] and more recently to distinguish between Alpha and Delta VOC in Scotland and England [2,9] and maybe highly useful when there is a large-scale testing strategy in place and electronic vaccination registers.

We cannot exclude delays in vaccination status and diagnosis registration. However, data extraction was performed three weeks after the end of the study period to minimize information bias due to misclassification of vaccination status and, for reference, vaccination centers are instructed to register vaccinations up to 24 hours after administration.

After adjusting for confounding, we did not observe a substantial change in OR estimates. Although we cannot exclude residual confounding bias, as other factors not accounted for might be associated with the probability of exposure to the virus and of being vaccinated (i.e. health and social care worker status, ethnicity, education). However, we found that other studies [9] that adjusted for these potential confounding factors, not accounted in our study, did not observe a substantial difference between crude and confounder-adjusted estimates. Hence, if any, residual confounding could have a small effect on the effect estimates.

Overall, we found significantly higher odds of vaccination in Delta cases when compared to Alpha cases, suggesting possible lower effectiveness of the mRNA vaccines in preventing infection with the Delta VOC. These findings can help decision-makers weigh on the application or lifting of control measures and adjusting vaccine roll-out depending on the predominance of the Delta variant and the coverage of partial and complete mRNA vaccination.

## Data Availability

Data will be available upon request to the Direcao Geral da Saude and Instituto Nacional de Saude Dr Ricardo Jorge

## Notes

**Conflict of interest:** None

### Competing Interest Statement

Eduardo Rodrigues (ED) board member and owns stocks at UPHILL, a software company that provides digital training solutions and that has customers inthe pharmaceutical sector (e.g. Pfizer). No business was conducted between the two entities regarding mRNA vaccine products or similar, neither relatedto the abovementioned study.

### Funding Statement

No funding is declared

### Author Declarations

This study was approved by Instituto Nacional de Saude Dr. Ricardo Jorge Ethical Committee

