## Supplementary material for "Delta variant and mRNA Covid-19 vaccines effectiveness: higher odds of vaccine infection breakthroughs"

**Figure S1 —** Vaccine uptake, from week 1 to 26 of 2021 in Mainland Portugal

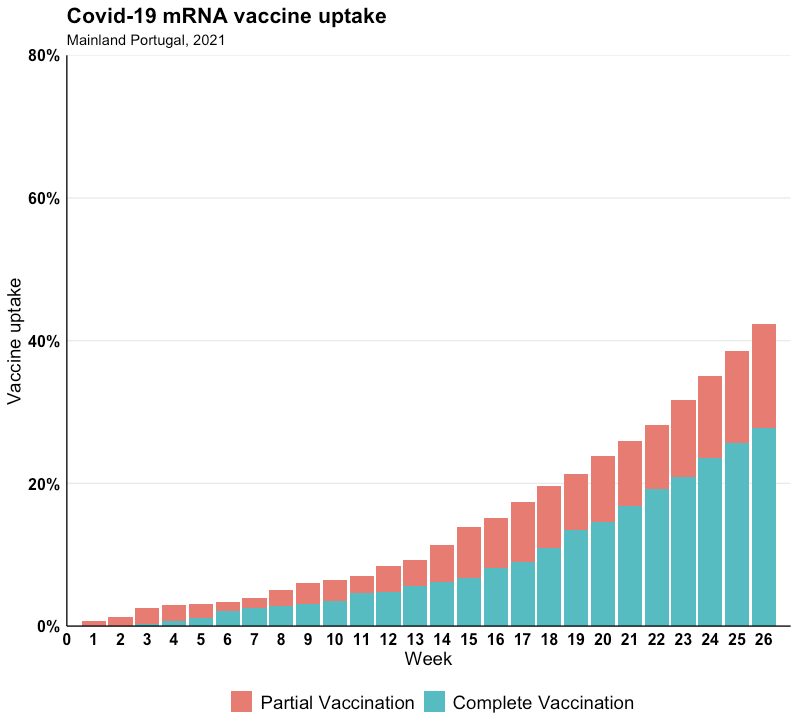

**Figure S2** — Number of people vaccinated with mRNA vaccine by brand, from week 1 to 26 of 2021 in Mainland Portugal

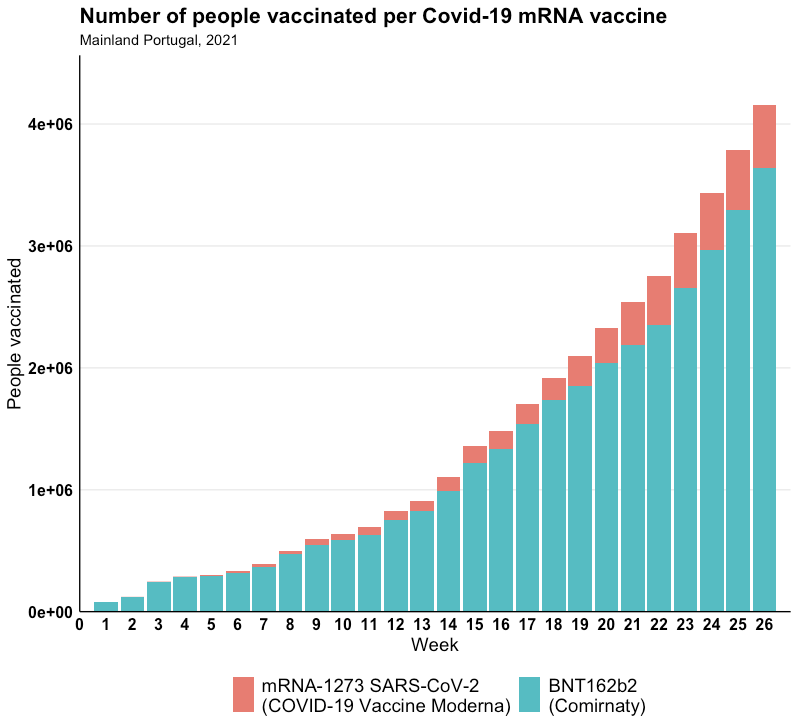

**Figure S3** — mRNA vaccine coverage per age group and week, from week 1 to 26 of 2021 in Mainland Portugal

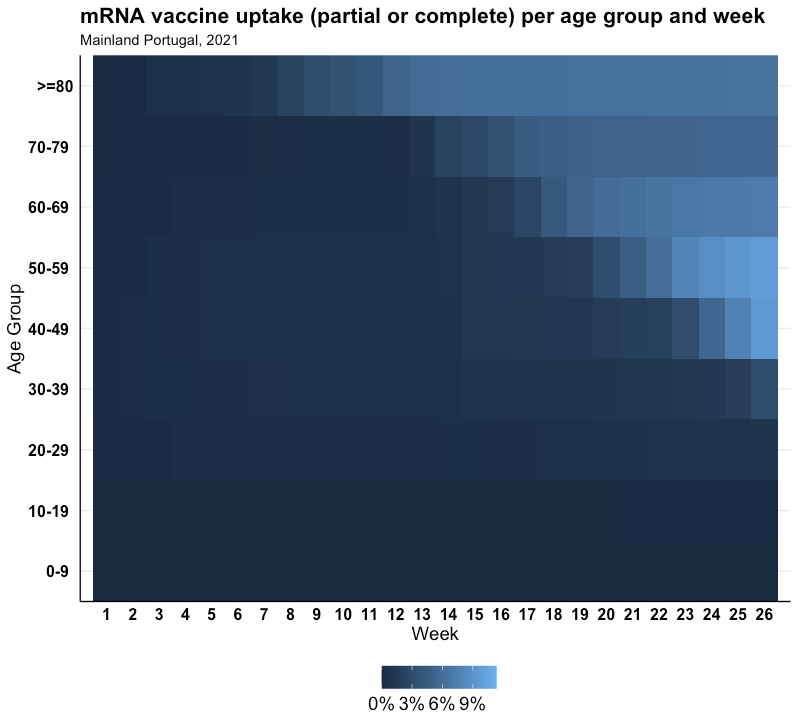

**Figure S4 —** Relative frequency of age groups of reported confirmed cases over 40 years to the national surveillance system and study sample, from week 20 to 26 of 2021 in Mainland Portugal

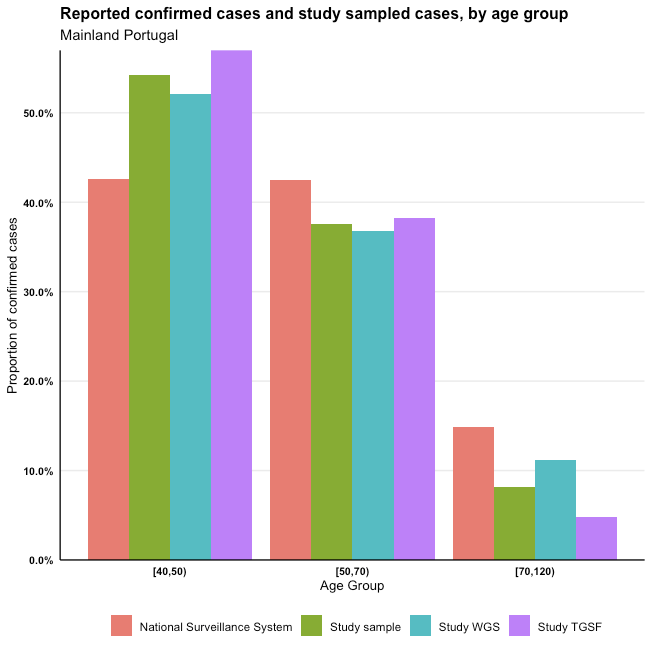

**Case-case study design description**

The most common approach of vaccine effectiveness (VE) studies is test-negative case-control design, where VE is estimated comparing odds of vaccination between positive (cases) and negative (controls) (VE=(1-OR)*100%).

Variant-specific vaccine effectiveness estimates in test-negative design (TND) studies are obtained restricting comparisons to cases positive to specific variant versus negative controls (Table S1)

**Table S1** — Classification of cases by vaccination and positivity status

|  | Delta+ | Alpha+ | Negative controls |
| --- | --- | --- | --- |
| Vaccinated | a | k | b |
| Unvaccinated | c | m | d |

resulting in vaccine effectiveness estimates

${VE}_{Delta}=\left( 1-{OR}_{Delta} \right)\times100\%$ and, ${VE}_{Alpha}=\left( 1-{OR}_{Alpha} \right)\times100\%$,

where ${OR}_{Delta}$ and ${OR}_{Alpha}$ are obtained as follows:

${OR}_{Delta}=\frac{a\times d}{c\times b}$ ${OR}_{Alpha}=\frac{k\times d}{m\times b}$

Only positive cases are considered in case-case design, and odds of vaccination are compared directly between two variants. Considering Delta cases as cases of interest and Alpha cases as a reference group, the case-case OR is estimated as:

$${OR}_{case-case}=\frac{a\times m}{c\times k}$$

that is mathematically equivalent to the ratio of two variant-specific OR obtained from TND:

$\frac{{OR}_{Delta}}{{OR}_{Alpha}}$=$\frac{a\times d}{c\times b}\div\frac{k\times d}{m\times b}$ =$\frac{a\times d\times m\times b}{c\times b\times k\times d}$=$\frac{a\times m}{c\times k}={OR}_{case-case}$

The case-case approach allows to formally test whether the difference between variant-specific odds ratios is statistically significant and infer variant-specific vaccine effectiveness (Figure S5).

**Figure S5** — Comparison diagram between test-negative design and case-case design

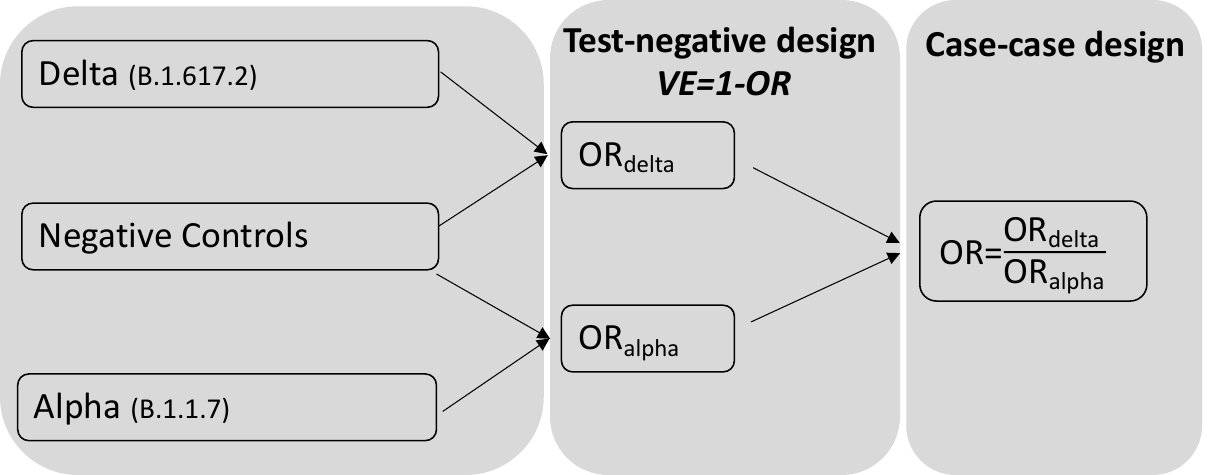
